# Using a large language model as the primary coder in health content analysis: a verification framework, per-variable reliability and proposed reporting items

**DOI:** 10.64898/2026.11.16.26368259

**Authors:** Isabelle Schlegel, Ailsa Sandilands, Gozde Ozakinci

## Abstract

**Objective:** To describe and evaluate a framework for using a large language model (LLM) as the primary coder in health content analysis, and to propose reporting items for LLM-assisted studies, because no reporting standard exists.

**Methods:** An LLM coded 6,982 Instagram and TikTok posts using a human-written, human-calibrated instrument. The framework has six components: human authorship of the instrument; two-round human calibration; source discipline; quote-anchored coding; verification in a representative and an outcome-enriched sample, blinded in both directions; and per-variable reliability reporting against a prespecified threshold, with the consequence of failure fixed in advance. Agreement was measured with Gwet’s AC1, prespecified because the outcome distribution was heavily skewed.

**Results:** Agreement between the two human coders on the primary outcome rose from AC1 0.456 to 0.977 after two ambiguous rules were rewritten. LLM–human agreement was 0.847 in the representative sample (N = 150) and 0.868 in the enriched sample (N = 50). Five of eight conditional variables fell below the 0.80 threshold and became exploratory: variables asking whether a defined textual feature was present met it, whereas those requiring graded judgement did not. The human coder identified misinformation more often than the LLM, so prevalence estimates are likely underestimates. Agreement on female targeting was lower for self-identified ethnic minority creators (0.744 versus 0.902).

**Conclusion:** LLM coding cannot be computationally reproduced, so its reliability must be shown empirically: per-variable agreement with independent human coding, the direction of remaining error, and a prespecified response to shortfall. The authors propose 18 reporting items for LLM-assisted content analysis.

## 1. Introduction

Content analysis of social media is commonly used to estimate how much of the information circulating on a platform is accurate, to characterise the forms that inaccurate information takes, and to identify the audiences it targets. Its value depends on scale: a prevalence estimate drawn from two hundred posts describes those posts, whereas one drawn from several thousand begins to describe an information environment.

Human coding does not reach that scale. Each post requires judgements about whether a claim is present, whether it is accurate and against what evidence, so a trained coder manages a few hundred posts a week. The literature reflects this constraint: a 2025 systematic review of physical activity misinformation found that most included studies examined YouTube or small TikTok samples [1]. The limitation is one of capacity, not rigour.

Large language models (LLMs) are being adopted to relieve this constraint, and adoption has outpaced reporting standards. Content analysis has no equivalent of STROBE or PRISMA. There is no agreed account of what readers need to be told to judge whether an LLM-coded corpus can be relied upon.

This paper describes a framework for making an LLM-coded study auditable, applies it to a corpus of 6,982 posts, reports its performance, including the variables on which it failed, and proposes reporting items for LLM-assisted content analysis.

### Statement of significance

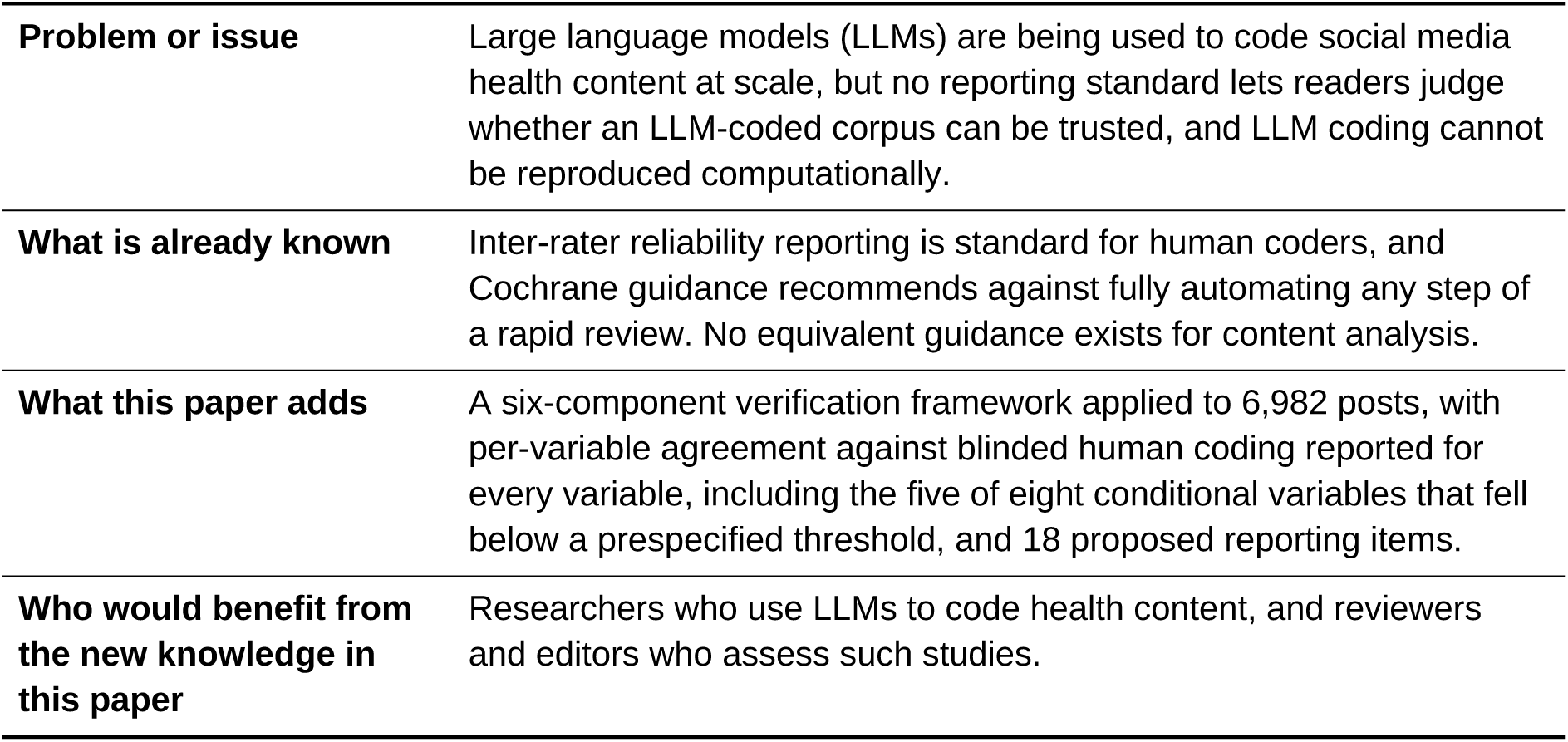

## 2. Related work

The nearest guidance concerns evidence synthesis. The Cochrane Rapid Reviews Methods Group recommends against fully automating any step of a rapid review, including screening, data extraction and risk-of-bias assessment [2], and the World Health Organization has issued guidance on ethics review and oversight of artificial intelligence (AI)-related health research [3]. Neither addresses content analysis, but the Cochrane position applies by extension. Coding in content analysis is analogous to data extraction: in both, a human judgement about a source document is converted into a structured variable.

The extension is not straightforward, because the alternative to automation differs. In a rapid review, the alternative to automated extraction is human extraction from a bounded set of studies, typically tens to low hundreds; the work is finite and automation saves time. In large-scale content analysis, the alternative is not slower human coding of the same corpus but a much smaller study, and a smaller study answers a different question. Where an effect is concentrated in a subgroup, or where a stratified design is needed to separate an exposure from an artefact of data collection, a corpus reduced fivefold may not answer the question at all.

This does not justify unverified automation. The pertinent question is not whether LLM coding matches human coding (on several variables reported below, it did not), but whether an LLM-coded study can be made auditable: whether a reader can see what the coder got right, what it got wrong, in which direction it erred and how the study approached this.

A second problem is that an LLM coder is not reproducible in the standard computational sense. The same prompt applied to the same posts need not return identical codes, and model versions are withdrawn and replaced faster than the publication cycle. A framework for this method therefore cannot promise reproducibility. What it can offer is a measured, per-variable account of agreement with independent human coding.

## 3. Methods

### 3.1 Setting

The framework was developed and applied within a cross-sectional content analysis of exercise misinformation in fitness content directed at women on Instagram and TikTok, which included a nested sub-study of content from creators who self-identified as an ethnic minority. The parent study collected 6,982 posts between March and June 2026 through two collection routes (manual and automated) and is reported separately [4]. This article concerns the coding method only.

The parent study was not pre-registered. A versioned protocol with dated amendments is deposited [5]. The reliability statistic, its threshold, and the primary analysis model were fixed before any results were seen.

### 3.2 The framework

The framework has six components, summarised in Fig. 1.

**Fig. 1.**
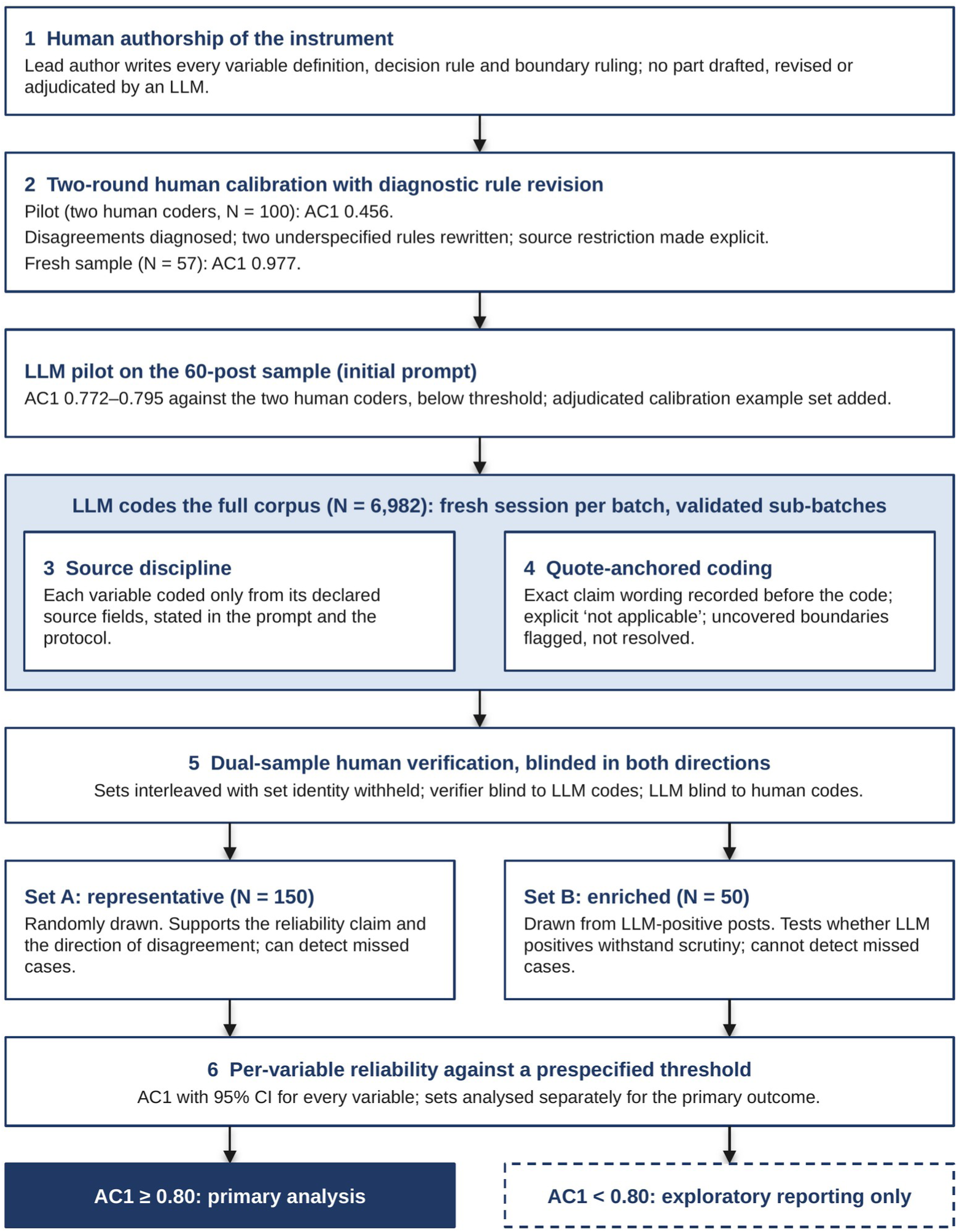
Framework for LLM-assisted content analysis with human verification. Numbered boxes are the six components of the framework. Components 3 and 4 are rules the LLM applied while coding the full corpus (shaded box). Set A (representative) supports the reliability claim for the dataset and the direction of disagreement; Set B (enriched for LLM-identified positives) tests whether those positives withstand human scrutiny but cannot detect missed cases. For the primary outcome the two sets are analysed separately. AC1, Gwet’s first-order agreement coefficient; CI, confidence interval; LLM, large language model.

#### Component 1: human authorship of the instrument

The lead author (IS) wrote the coding instrument before any LLM-assisted coding. No variable definition, decision rule or boundary ruling was drafted, revised or adjudicated by an LLM at any point. The standard against which the LLM coder is judged is therefore a human one, and a failure to meet it on a given variable is interpretable rather than circular.

#### Component 2: two-round human calibration with diagnostic rule revision

The instrument was tested by two human coders: the lead author (IS) and a second coder (AS), who had no part in writing it. Both coded a calibration sample independently, and agreement was computed. Disagreements were treated as diagnostic; each was examined to establish whether it arose from inconsistent application of a rule that determined an answer, or from a rule that did not. In the latter case, the rule was rewritten, and agreement was recomputed on a fresh sample, following the iterative codebook development described by Hruschka et al. [6]. Both the initial and final figures are reported.

#### Component 3: source discipline

Each variable could be coded only from its declared source fields, and this restriction was stated in the coding prompt as well as in the protocol. The primary outcome was coded from caption and on-screen text only, never from hashtags or creator biographies; professional status from biography text only; and ethnic minority self-identification from explicit self-statements in text only. No variable was coded from images, video, or usernames. This rule was adopted first because inferring ethnicity from photographs or names is ethically indefensible, and was then applied to every variable for consistency.

#### Component 4: quote-anchored coding

The exact wording of the claim on which a code rested had to be recorded in a notes field before the code was assigned, so that the textual basis for each decision preceded the judgement rather than being reconstructed afterwards. Every coding decision can therefore be traced to, and checked against, the text that produced it. An explicit “not applicable” code was required instead of a blank, because a blank conditional variable is not a zero; recording no rhetorical features for a post without a claim asserts something the coder did not assess. Where the rules did not cover a case, the coder applied the nearest written rule, recorded the alternative code considered and set an ambiguity flag, rather than resolving the boundary. Resolving new boundaries partway through a sample changes the instrument mid-sample and invalidates reliability computed across the whole.

#### Component 5: dual-sample human verification with two-sided blinding

The LLM coded the full corpus in a fresh session for each batch, working in sub-batches with a validation check after each. Its coding was then verified against human coding in two samples. For the primary outcome, the two samples were analysed separately and never combined:

- Set A, drawn to be representative of the corpus (N = 150). This sample supports the reliability claim for the dataset and is the only one of the two that can detect misinformation the LLM missed.
- Set B, enriched for the outcome (N = 50). Set B was drawn from posts the LLM had coded as containing misinformation (Yes or Mixed); it assesses whether LLM-identified positives withstand human scrutiny. It cannot detect false negatives and should not be read as a general agreement estimate.

Blinding operated in both directions. The LLM was blinded to all human codes: every reliability or verification sample was coded in a session containing no human codes, reliability results or reconciliation discussion, because exposure to any of these would invalidate the estimate. The verifying human coder received Sets A and B interleaved in random order, with the set identifier withheld, and coded without sight of the LLM codes.

Conditional variables are defined only for posts coded as containing misinformation, so they were assessed on the posts that both coders coded as misinformation across Sets A and B (n = 57); Set B supplied most of these posts. Female targeting was also estimated across both sets, as a secondary estimate (see Section 4.5).

The authors considered and rejected using the LLM to pre-filter posts before human coding (for example, removing posts it judged to contain no assessable claim). Pre-filtering would be circular, because it would conceal the coder’s errors at exactly the judgement boundary where they are most likely.

#### Component 6: per-variable reliability reporting against a prespecified threshold

Agreement was computed and reported separately for every coded variable, including those that fell short. A threshold of AC1 ≥ 0.80 was fixed in advance and applied to the point estimate, with the confidence interval reported. Variables below the threshold were moved to exploratory reporting, and analyses resting on them were reported as exploratory or withdrawn. The consequence of failure was therefore set before the results were known.

### 3.3 Model and configuration

Coding used Claude Opus 5 (Anthropic) at high reasoning effort, accessed through the Claude app with the setting that allows conversations to be used for model training switched off, between late May and late August 2026. Each batch was coded in a fresh session rather than a continuing one, to prevent context from earlier batches influencing later ones and to enforce the blinding rule above.

Because LLM outputs can vary between runs and model versions are withdrawn, computational reproducibility is not claimed for this coder. The claim made instead is empirical agreement with independent human coding, reported for each variable. The full prompt scaffold is deposited with the machine-readable codebook [5]. In summary, the scaffold supplied the protocol, the log of boundary rulings, the coding quick reference and the adjudicated calibration examples; stated the source restriction for each variable; required the exact claim wording before each code; required an explicit “not applicable” code; instructed the coder to flag rather than resolve uncovered boundaries; and coded in validated sub-batches.

Where the protocol text and the adjudicated calibration examples appeared to conflict, the calibration examples took precedence, because they record decisions already made about those boundary cases.

### 3.4 Reliability statistic

Gwet’s AC1 [7] was prespecified as the primary agreement statistic and fixed before results were seen, to prevent post hoc selection. The reason is the marginal distribution of the outcome. Where one category predominates (many posts contained no assessable claim in this sample), Cohen’s kappa can return low values despite high observed agreement, because its chance correction assumes coders guess in proportion to the observed marginal proportions. This is the kappa paradox: a property of skewed marginals rather than poor coding.

The difference is material at the levels relevant to this design. For creator professional status in Set A, 85.3% observed agreement gave AC1 0.825 but kappa 0.550; in the first human calibration round, 58.0% observed agreement gave AC1 0.456 and kappa 0.408.

Cohen’s kappa was computed alongside AC1 throughout and is reported in the deposited materials for comparison with earlier studies. AC1 variance was estimated with the linearisation estimator [7]. The AC1 implementation was written for this study, validated against published figures and deposited [5].

### 3.5 Subgroup performance

Because LLMs are trained predominantly on sources from high-income, predominantly English-speaking countries, poorer performance on content from minority creators is plausible and has been raised in guidance on AI in health research [3]. The authors tested this rather than assuming it in either direction, by computing agreement separately for posts from creators who self-identified as an ethnic minority and for all other posts. The general rule the authors followed is to stratify the reliability sample on any subgroup variable that carries an inferential claim.

A post was placed in the subgroup if either coder flagged the creator as self-identifying as an ethnic minority, and in the comparison group if neither did; agreement was computed across Sets A and B for the primary outcome and for female targeting.

### 3.6 Independent reimplementation

All analyses were conducted in jamovi 2.7.37 [8] and independently reimplemented in Python (pandas, SciPy, statsmodels), with exact agreement required before any result was reported.

### 3.7 Open materials

The coded dataset, annotated analysis script, machine-readable codebook with per-variable reliability, prompt scaffold, causal model, AC1 implementation and versioned protocol are deposited [5]. The public dataset is anonymised and contains no raw platform text, creator handles, exact dates or engagement counts.

## 4. Results

### 4.1 Reliability by variable

Agreement for every coded variable is shown in Table 1 and Fig. 2.

**Table 1.** Inter-rater reliability (Gwet’s AC1) by variable and comparison.

| Variable | Comparison | N | AC1 | 95% CI | ≥ 0.80 |
|---|---|---|---|---|---|
| **Instrument calibration, primary outcome (human–human)** |  |  |  |  |  |
| Misinformation | IS vs AS, pilot<sup>a</sup> | 100 | 0.456 | – | – |
| Misinformation | IS vs AS, after rule revision | 57 | 0.977 | 0.931–1.000 | Yes |
| **LLM pilot, initial prompt (60-post sample)**<sup>b</sup> |  |  |  |  |  |
| Misinformation | LLM vs IS | 57 | 0.772 | 0.634–0.909 | – |
| Misinformation | LLM vs AS | 57 | 0.795 | 0.665–0.925 | – |
| **Verification: primary outcome** |  |  |  |  |  |
| Misinformation | LLM vs AS, Set A (representative) | 150 | 0.847 | 0.781–0.914 | Yes |
| Misinformation | LLM vs AS, Set B (enriched)<sup>c</sup> | 50 | 0.868 | 0.756–0.980 | Yes |
| **Verification: other universal variables (Set A)** |  |  |  |  |  |
| Female targeting (three levels) | LLM vs AS | 150 | 0.817 | 0.745–0.889 | Yes |
| Ethnic minority self-identification | LLM vs AS | 150 | 0.867 | 0.802–0.932 | Yes |
| Professional status | LLM vs AS | 150 | 0.825 | 0.754–0.897 | Yes |
| Monetisation | LLM vs AS | 150 | 0.853 | 0.785–0.922 | Yes |
| Quality score (weighted AC2) | LLM vs AS | 46 | 0.923 | 0.886–0.960 | Yes |
| Exercise topic (five levels) | LLM vs AS | 150 | 0.782 | 0.709–0.855 | No |
| **Verification: conditional variables (Sets A and B)**<sup>d</sup> |  |  |  |  |  |
| Misuse of scientific reference | LLM vs AS | 57 | 0.960 | 0.903–1.000 | Yes |
| Pseudo-scientific language | LLM vs AS | 57 | 0.904 | 0.809–1.000 | Yes |
| Quick fix | LLM vs AS | 57 | 0.870 | 0.762–0.978 | Yes |
| Number of misinformation categories | LLM vs AS | 57 | 0.613 | 0.448–0.779 | No |
| Misinformation category (six levels) | LLM vs AS | 57 | 0.609 | 0.464–0.754 | No |
| Absolute claim | LLM vs AS | 57 | 0.479 | 0.250–0.709 | No |
| Commercial prompt | LLM vs AS | 57 | 0.404 | 0.167–0.642 | No |
| Guaranteed results | LLM vs AS | 57 | 0.378 | 0.135–0.621 | No |
| **Post hoc contrasts and secondary estimates** |  |  |  |  |  |
| Misinformation category, aesthetic vs other<sup>e</sup> | LLM vs AS, Sets A and B | 57 | 0.803 | 0.650–0.956 | Yes |
| Female targeting (three levels)<sup>f</sup> | LLM vs AS, Sets A and B | 200 | 0.847 | 0.786–0.909 | Yes |
| Female targeting, any vs none<sup>e,f</sup> | LLM vs AS, Sets A and B | 200 | 0.911 | 0.857–0.966 | Yes |

AC1, Gwet's first-order agreement coefficient; AC2, Gwet's weighted agreement coefficient (quadratic weights); AS, second human coder and verifier; CI, confidence interval; IS, lead author; LLM, large language model. The threshold of AC1 ≥ 0.80 was prespecified for every variable; variables below it are reported as exploratory. Confidence intervals use Gwet's linearisation variance, truncated at 1. The ambiguity flag, a process measure rather than a reliability outcome, had AC1 0.662 in Set A.
<sup>a</sup> Superseded: two coding rules were rewritten after this round; no confidence interval was computed for this round.
<sup>b</sup> Blind LLM coding with the initial prompt scaffold, before the calibration example set was added. Superseded.
<sup>c</sup> Drawn from posts the LLM coded as containing misinformation; tests LLM-identified positives only and cannot detect false negatives.
<sup>d</sup> Defined only for posts coded as containing misinformation; assessed on the 57 posts that both coders coded as misinformation.
<sup>e</sup> Post hoc collapse to the binary contrast used in analysis.
<sup>f</sup> Secondary estimate. Set B was selected on the LLM's misinformation code, not its targeting code; the Set A estimate carries the reliability claim. Non-English posts coded N/A are counted as not targeted.

**Fig. 2.**
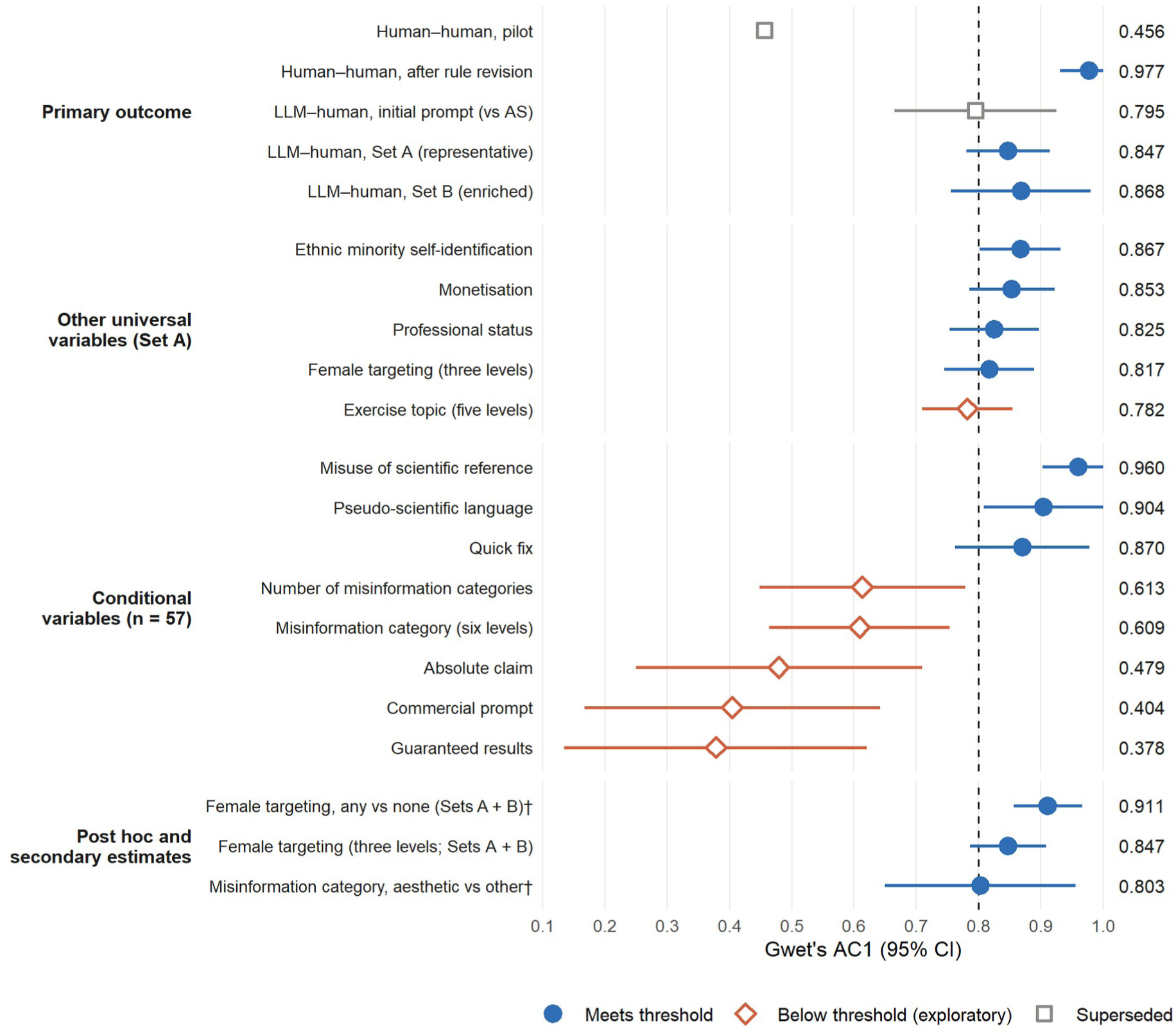
Agreement by variable against the prespecified threshold. Points show Gwet’s AC1 with 95% confidence intervals; the dashed line marks the prespecified threshold of 0.80. Filled circles met the threshold; open diamonds fell below it and were moved to exploratory reporting; open squares are superseded rounds (the human pilot, before two coding rules were rewritten, and the LLM pilot with the initial prompt). No interval is shown for the human pilot. Other universal variables are from Set A (N = 150); conditional variables from the 57 posts both coders coded as misinformation across Sets A and B. † Post hoc collapse to the binary contrast used in analysis. Quality score (weighted AC2 0.923) is reported in Table 1. LLM, large language model; AS, second human coder.

### 4.2 Instrument calibration

At first calibration (N = 100), agreement between the two human coders on the primary outcome was AC1 0.456. Examination of the disagreements showed that they arose from two rules that did not determine an answer in the disputed cases, rather than from inconsistent application of rules that did. The largest single cause was source leakage; in some cases, the primary outcome was being coded from hashtags rather than from caption and on-screen text, producing systematic false positives. Both rules were rewritten, and the source restriction was made explicit for each variable. On a fresh sample of 60 posts (57 after all coders excluded the same three non-English posts), the same two coders reached AC1 0.977 (95% CI 0.931 to 1.000). The human pair met the threshold on every variable except exercise topic (0.795, 95% CI 0.671 to 0.920).

This interpretation is crucial to the logic of the framework. If the first figure had been read as evidence of unreliable coders, the response would have been retraining, and the ambiguity in the rules would have carried into the full corpus. There it would have been inherited by the LLM coder, which applies a written rule far more consistently than a human does and would therefore have propagated the ambiguity uniformly and invisibly. Low initial agreement between human coders is more usefully read as a property of the instrument than of the coders.

### 4.3 LLM pilot

Before the full corpus was coded, the LLM coded the same 60-post sample blind, using the initial prompt scaffold. Agreement on the primary outcome was below the threshold: 0.772 (95% CI 0.634 to 0.909) against IS and 0.795 (0.665 to 0.925) against AS, falling to 0.457 and 0.533 on the 19 posts carrying a claim. The LLM coded Mixed less often than either human. The adjudicated calibration example set was built to address this boundary, and the full corpus was coded with it. The pilot figures are superseded and are reported for completeness; the verification below applies to the final coding.

### 4.4 LLM verification

Agreement between the LLM and the human verifier on the primary outcome exceeded the threshold in the representative sample (Set A, AC1 0.847, 95% CI 0.781 to 0.914). Set B returned 0.868 (0.756 to 0.980), indicating that LLM-identified positives largely withstood human scrutiny; by design, it carries no information about missed cases. In Set A, the other universal variables met the threshold: female targeting (0.817), ethnic minority self-identification (0.867), professional status (0.825), monetisation (0.853) and quality score (weighted AC2 0.923; n = 46). Exercise topic did not (0.782, 95% CI 0.709 to 0.855).

Of the eight conditional variables, assessed on the 57 posts that both coders coded as misinformation, three met the threshold: misuse of scientific reference (0.960), pseudo-scientific language (0.904) and quick fix (0.870). Five did not: number of misinformation categories (0.613), misinformation category at six levels (0.609), absolute claim (0.479), commercial prompt (0.404) and guaranteed results (0.378). Confidence intervals for every variable are given in Table 1.

### 4.5 Reliability at the level of analysis

Two variables were collapsed post hoc to the binary distinction that their analyses actually test; both are labelled as post hoc throughout. Misinformation category rose from 0.609 at six levels to 0.803 (95% CI 0.650 to 0.956) for aesthetic claims versus all other categories. Female targeting was examined as any targeting versus none.

Female targeting is also reported across Sets A and B combined (N = 200) as a secondary estimate. Set B was selected on the LLM’s misinformation code, not its targeting code, so combining the samples adds precision without conditioning on the variable being assessed; the Set A estimate (0.817) carries the reliability claim. Combined, agreement was 0.847 (95% CI 0.786 to 0.909) at three levels and 0.911 (0.857 to 0.966; 95.0% observed agreement) for any targeting versus none, with the non-English posts coded N/A counted as not targeted.

The collapse is informative, not merely convenient. For female targeting, disagreement arose almost entirely from the LLM classifying implicit targeting as explicit (12 of 30 posts coded as implicit by the human coder), whereas the explicit and no-targeting categories almost perfectly agreed. The level at which a variable is reliable therefore depends on the boundary being drawn. A multi-level variable can sit near the threshold overall, while the individual levels a study utilises for detailed analysis sit comfortably above it. Reporting reliability at the level of analysis, rather than the level of the codebook, avoids both discarding a usable measure and trusting an unusable distinction.

### 4.6 Variables coded reliably by the LLM

The pattern of success and failure is interpretable rather than random, and is the most transferable empirical finding of this study. The variables that met the threshold ask whether a specified textual feature is present: a reference to science, pseudo-scientific vocabulary, a promise of speed. The variables that fell short ask for a graded judgement about persuasive intent, or about which of several competing categories is most salient: whether a claim is absolute rather than merely strong, whether a post prompts a commercial action rather than merely mentioning a product, or which of six misinformation categories predominates. The LLM handled feature detection well and graded judgement poorly.

This has a direct implication for instrument design in LLM-assisted content analysis. Variables should be specified as feature-presence tests wherever the construct allows, and constructs that unavoidably require graded judgement should be expected to need human coding or to be reported as exploratory. Female targeting illustrates both points: whether a gendered term of address is present is a feature-presence test and was coded reliably, whereas whether a post is directed at women without saying so is a graded, inferential judgement and was not. The pattern is visible only because failing variables were reported, alongside the collapsed contrasts.

### 4.7 Consequences of failure

The five conditional variables below the threshold were moved to exploratory reporting under the rule fixed in advance. In the parent study, this affected two of four secondary aims: the analyses of misinformation categories and of rhetorical features are reported as exploratory, and the distribution of misinformation categories is explicitly not reported as a measured prevalence. Exercise topic, a universal variable, also fell below the threshold in Set A (0.782) and is used in the parent study only as an adjustment covariate and in descriptive comparisons, which are labelled exploratory.

### 4.8 Direction of disagreement

Disagreement on the primary outcome was not symmetrical. In Set A, the human coder coded 21 posts as containing misinformation and the LLM 10. All 11 disagreements about whether misinformation was present ran the same way: the human identified misinformation that the LLM did not. Remaining measurement error therefore runs towards under-detection, and prevalence estimates in the parent study are likely to be underestimates rather than estimates with error of unknown direction.

This could be reported only because the verification design facilitated it. An agreement statistic alone has no direction: AC1 0.847 is consistent with the LLM either over-calling or under-calling, and the two have opposite consequences for how a prevalence estimate should be read. It can also be reported only from Set A, which is why the two samples must be analysed separately.

### 4.9 Subgroup performance

Agreement on the primary outcome was similar for posts from creators who self-identified as an ethnic minority (AC1 0.848, 95% CI 0.748 to 0.949; n = 65) and for other posts (0.833, 0.756 to 0.910; n = 126). Agreement on female targeting was not: 0.744 (0.609 to 0.879) in the subgroup, below the threshold, against 0.902 (0.839 to 0.965) for other posts. The shortfall lay in separating implicit from explicit targeting. Of the 11 subgroup posts that the human coder coded as implicitly targeted, the LLM coded 8 as explicit and 2 as implicit, whereas agreement on any targeting versus none held (0.889, 0.780 to 0.997). The split between implicit and explicit targeting among ethnic minority creators in the parent study is therefore not reliably measured. The LLM also under-detected self-disclosed minority identity: of 65 posts the human coder flagged, the LLM coded 15 as not self-identifying, so the flagged subgroup is conservative. The test covers only creators who disclosed an identity in text, and does not address performance on content from minority creators in general.

### 4.10 Independent reimplementation

Reimplementation reproduced every reported result exactly, with one exception. The first Python implementation of the primary regression excluded rows with a missing value on any predictor (n = 2,365), whereas the jamovi implementation retained missingness as a separate category (n = 2,447). Both were arithmetically correct, but odds ratios differed by up to 5%.

The divergence was traced to an unstated analytical decision rather than to computation and was resolved by adopting one convention explicitly in both environments (complete-case analysis, excluding posts coded N/A on a model predictor). This failure mode is worth highlighting because it is the one that dual implementation identifies: not arithmetic error, but a specification choice that neither implementation had been required to state. It has the same structure as the instrument failure in the first calibration round: an underspecified rule, applied consistently but differently by two competent agents.

## 5. Discussion

### 5.1 Principal contribution

The framework does not make LLM coding equivalent to human coding; on five of eight conditional variables it was clearly not. What it does is make the limitations visible, bounded and consequential: visible because agreement is reported for every variable, including those that fall short; bounded because the direction of remaining error on the primary outcome is known; and consequential because the handling of failing variables was fixed before the results were seen.

This is a lower bar than validity, but a more achievable one. A reader of a study reported in this way does not have to trust the coder. They can see which variables to trust, which to discount, and in which direction the remaining error runs.

### 5.2 Reproducibility, and what replaces it

An LLM coder is not directly reproducible. Identical inputs need not return identical outputs; models are versioned, deprecated and withdrawn faster than papers are published; and a study coded on one model version cannot be rerun on it indefinitely. A framework that implied otherwise would promise something it cannot deliver.

What can be offered instead is a measured agreement claim for each variable against independent human coding, together with the deposited instrument, prompt scaffold and codebook. These allow another team to apply the same instrument with a different coder, human, or model, and compare the results. This is a weaker claim than computational reproducibility, but a stronger one than is currently available.

### 5.3 Relation to existing guidance

The Cochrane position that no step of a rapid review should be fully automated [2] is not contradicted here, because no step was fully automated. The instrument was written by a human, verification was carried out by a human, and the rule for handling failure was set by the authors in advance. The LLM applied a human-written instrument at a scale humans cannot reach, under human audit.

Where this analysis diverges from a strict reading is in the implied comparison. The Cochrane statement itself cites evidence of substantial error in human data extraction, with rates of up to 50% [9], and the human coders here began at AC1 0.456 on the primary outcome. Treating human coding as the reliable baseline and LLM coding as a deviation from it describes neither accurately. The realistic comparison is between an LLM-coded corpus of 6,982 posts with per-variable verification and a human-coded corpus of 1,200 posts with reliability reported on a subsample. The second is not confidently more reliable, and it does not answer the same question.

### 5.4 Proposed reporting items

Table 2 lists items the authors suggest should be reported in any content analysis that uses an LLM as a coder. They derive from the framework above and from the failure modes it exposed. The list has not been validated and is offered as a starting point for consensus work, not as a standard.

**Table 2.**
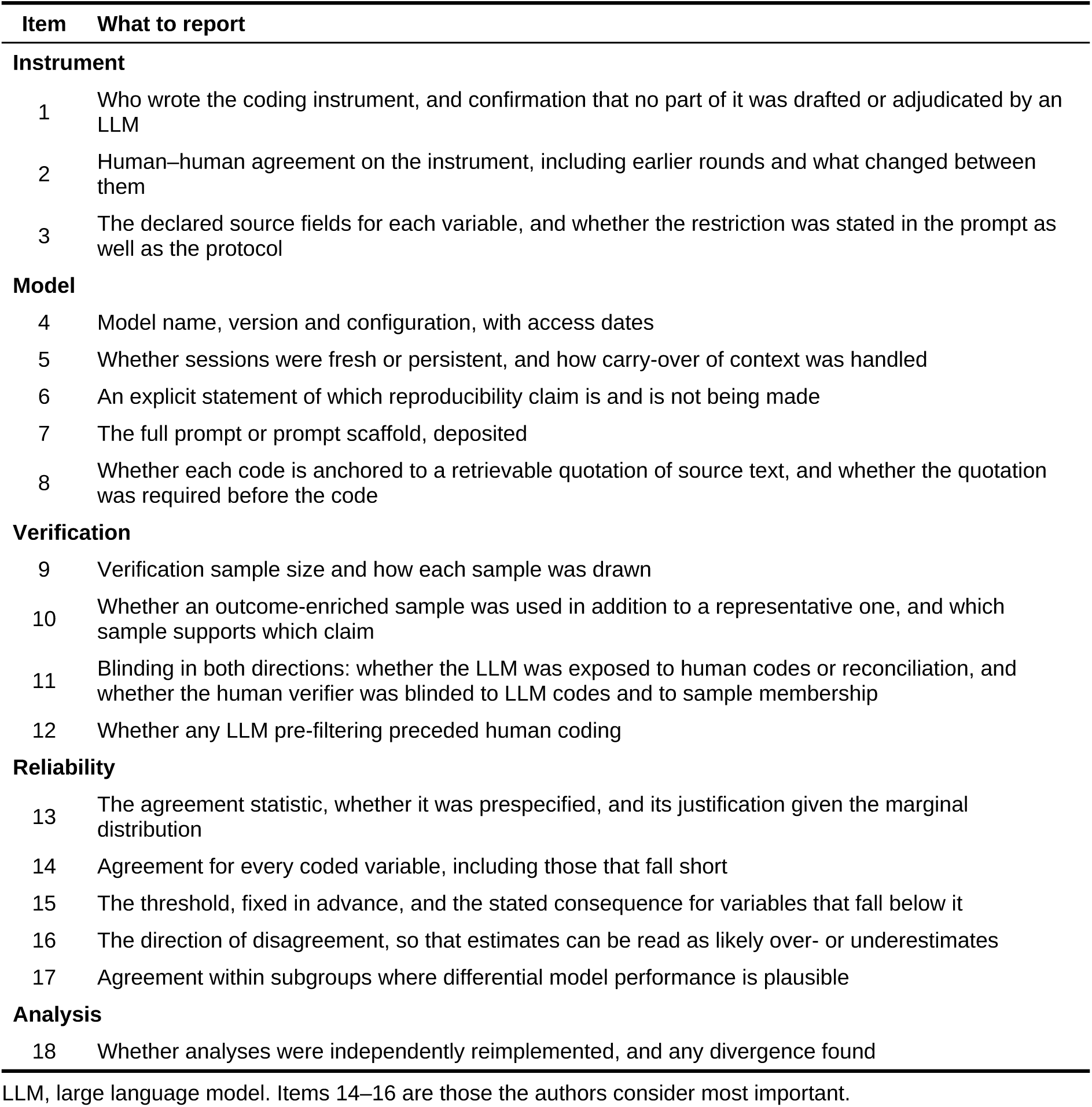
Proposed reporting items for LLM-assisted content analysis.

| Item | What to report |
|---|---|
| **Instrument** |  |
| 1 | Who wrote the coding instrument, and confirmation that no part of it was drafted or adjudicated by an LLM |
| 2 | Human–human agreement on the instrument, including earlier rounds and what changed between them |
| 3 | The declared source fields for each variable, and whether the restriction was stated in the prompt as well as the protocol |
| **Model** |  |
| 4 | Model name, version and configuration, with access dates |
| 5 | Whether sessions were fresh or persistent, and how carry-over of context was handled |
| 6 | An explicit statement of which reproducibility claim is and is not being made |
| 7 | The full prompt or prompt scaffold, deposited |
| 8 | Whether each code is anchored to a retrievable quotation of source text, and whether the quotation was required before the code |
| **Verification** |  |
| 9 | Verification sample size and how each sample was drawn |
| 10 | Whether an outcome-enriched sample was used in addition to a representative one, and which sample supports which claim |
| 11 | Blinding in both directions: whether the LLM was exposed to human codes or reconciliation, and whether the human verifier was blinded to LLM codes and to sample membership |
| 12 | Whether any LLM pre-filtering preceded human coding |
| **Reliability** |  |
| 13 | The agreement statistic, whether it was prespecified, and its justification given the marginal distribution |
| 14 | Agreement for every coded variable, including those that fall short |
| 15 | The threshold, fixed in advance, and the stated consequence for variables that fall below it |
| 16 | The direction of disagreement, so that estimates can be read as likely over- or underestimates |
| 17 | Agreement within subgroups where differential model performance is plausible |
| **Analysis** |  |
| 18 | Whether analyses were independently reimplemented, and any divergence found |

LLM, large language model. Items 14–16 are those the authors consider most important.

Items 14, 15 and 16 carry the greatest significance. Reporting only the variables that met the threshold, reporting agreement without stating what would have happened had it been lower, or reporting a direction-free statistic for an estimate that will be read as a prevalence each leaves readers unable to judge how far the results can be trusted.

### 5.5 Scope of the framework

The framework does not establish that the LLM’s codes are correct, only that they agree with a human coder’s to a measured degree in a sample. Agreement is not accuracy; both coders may be wrong in the same direction against an external standard. It offers no remedy for the variables the coder could not handle; graded judgements about persuasive intent were lacking here, and the framework detects the failure without rectifying this. It is also restricted to written text. Content carried in images, video or audio was outside the coding scheme, which limits what any reliability figure reported here can signify.

### 5.6 Limitations

The verification samples are modest (N = 150 and N = 50), and the figures for conditional variables rest on 57 posts. Confidence intervals around AC1 at these sample sizes are wide, so the ordering of variables above and below the threshold should not be over-interpreted; the threshold crossings are the interpretable result.

The second coder did not write the instrument and coded the verification samples blind to the LLM codes and to sample membership, but they were not independent of the instrument’s development. Their disagreements at first calibration identified the two underspecified rules, and the instrument was revised in response. This is the iterative method working as intended [6], not a methodological error, but it limits what the final figure signifies. AC1 0.977 should be read as agreement reached by the development pair on an instrument refined using disagreements, not as an estimate of what the deposited instrument would achieve with a new coder. The latter is the quantity relevant to transferability, and this study does not measure it. The same coder also provided both the human–human and the LLM–human comparisons, so those figures are not independent of each other.

The subgroup analysis captures only creators who disclosed an ethnic minority identity in text. This conservative rule under-identifies the subgroup and leaves performance on content from undisclosed minority creators untested.

The framework was developed and applied within a single study, in one content domain, with one model. Its transferability to other settings is untested.

## 6. Conclusions

An LLM can serve as the primary coder in a health content analysis at a scale human coding cannot reach, and the resulting study can be made auditable. The conditions are that the instrument is written by humans before coding is automated; that verification is designed to reveal the direction of error, not only its size; that reliability is reported for every variable, including those that fall short; and that the consequence of falling short is fixed before results are known.

Computational reproducibility is not available and should not be claimed. Reported as described here, an LLM-coded study asks readers to check rather than to trust. That is a reasonable request, and the current absence of a reporting standard makes it hard to meet.

The framework applies to studies in which an LLM codes written text in health content, such as post captions and on-screen text, and a human-written instrument and an independent human verifier are available. It has not been tested on image, video or audio content, with other models, or in other content domains.

## Data availability

The datasets generated and analysed during the current study are available in the Zenodo repository, https://doi.org/10.5281/zenodo.22867157 [5]. The deposit comprises the coded dataset, analysis script, machine-readable codebook with per-variable reliability, prompt scaffold, causal model, AC1 implementation and versioned protocol. The dataset is anonymised for public release and contains no raw platform text, creator handles, exact dates or engagement counts.

## Ethics statement

The parent study was reviewed by the University of Stirling ethics panel and confirmed on 30 January 2026 as exempt from full ethics review through the low-risk checklist route (reference EC 2026 26091 20102). The study is exempt because it analysed publicly available social media posts and recruited, contacted and intervened with no participants. Rights and privacy were protected by pseudonymising creator and post identifiers, withholding raw platform text, creator handles, exact dates and engagement counts from the public dataset, and quoting no post verbatim. Data were processed under Articles 6(1)(e) and 9(2)(j) of the UK General Data Protection Regulation. A subsequent change request was approved by the same panel on 13 November 2026 (reference EC 2026 26091 22574).

## CRediT authorship contribution statement

**Isabelle Schlegel:** Conceptualization, Methodology, Investigation, Formal analysis, Writing – original draft. **Ailsa Sandilands:** Investigation, Validation. **Gozde Ozakinci:** Supervision, Writing – review and editing.

## Declaration of generative AI and AI-assisted technologies in the manuscript preparation process

During the preparation of this work the authors used Claude (Anthropic) to edit and tighten the text, to check reported figures against the study’s results files, and to format the manuscript and tables. After using this tool, the authors reviewed and edited the content as needed and take full responsibility for the content of the published article. The use of an LLM as the primary coder is the subject of this study and is described in Section 3. No AI tool is listed as an author.

## Acknowledgements

The authors thank BASEM for supporting this work through the Dr Vivien Lane Scholarship.

